# Characteristics and Prognosis of HHV8-Positive Diffuse Large B-Cell Lymphoma, Not Otherwise Specified: A Population-Based Study

**DOI:** 10.1101/2023.07.17.23292761

**Authors:** Fan Wang

## Abstract

**Background:** HHV8-positive diffuse large B-cell lymphoma, not otherwise specified (HDN) is a rare and aggressive lymphoma that usually arises in association with HHV8-positive multicentric Castleman disease. The epidemiology, treatment patterns and survival outcomes of HDN are poorly understood.

**Methods:** A retrospective analysis was performed for 67 patients with HDN diagnosed from 2011 to 2020 using the SEER database. The clinicopathologic characteristics, treatment modalities and survival outcomes of HDN patients were evaluated. Kaplan-Meier analysis and Cox regression analysis were employed to identify prognostic factors for overall survival (OS) and disease-specific survival (DSS).

**Results:** The median age at diagnosis was 51.8 years, and 79.1% of patients were male. The primary site distribution was mainly nodal (79.1%), while the extranodal sites were rarely involved (20.9%). The majority of patients were white (65.7%). Only 3.0% of patients received radiotherapy, while 55.2% received chemotherapy. The 1-year, 3-year and 5-year OS was 67.4%, 65.6%, 58.4%, and 56.3%, respectively, and the corresponding DSS was 73.1%, 73.1%, and 67.8%, respectively. The diagnosis year group of 2016-2020 had a significantly worse OS than the diagnosis year group of 2011-2015 (P = 0.040), but not for DSS (P = 0.074). No significant survival improvement was observed in patients underwent chemotherapy. Age and marital status were independent prognostic factors for OS, and age was an independent prognostic factor for DSS.

**Conclusions:** HDN is a rare and aggressive disease, our study provides a comprehensive overview of the epidemiology, treatment patterns and survival outcomes of HDN patients for the first time. We revealed that older age and marital status of single were associated with worse survival of HDN, while chemotherapy was not associated with improved survival outcomes in HDN patients.

## Introduction

Human herpesvirus 8 (HHV8), also known as Kaposi sarcoma-associated herpesvirus (KSHV), is a DNA virus with the ability to infect various cell types, leading to the development of different diseases, particularly in individuals with compromised immune systems^1^. One of the diseases associated with HHV8 infection is HHV8-positive diffuse large B-cell lymphoma, not otherwise specified (HHV8+ DLBCL-NOS or HDN). This is a rare and aggressive subtype of large B-cell lymphoma that is commonly linked to HHV8-positive multicentric Castleman disease (MCD), a disorder characterized by systemic inflammation and enlargement of lymph nodes^2–5^. However, some cases of HDN were also found in the absence of MCD^6, 7^. The incidence of HDN is a quite low, with about 0.1%^8^. It is predominantly observed in patients with HIV, but it can also arise in other situations of immunodeficiency or immunosuppression^9^. The clinical presentation of HDN is variable, and these neoplasms usually involve lymph nodes^3^. However, it can also spread to extranodal sites and cause significant splenomegaly with peripheral blood involvement^3^. In rare cases, these neoplasms may be limited to specific organs such as the heart, spleen, or liver^10–12^.

HDN is characterized by a diffuse and abnormal growth of large lymphoid cells infected with HHV8, which exhibit a resemblance to plasma blasts and express IgM lambda^3, 13^. The tumor cells typically show varying levels of CD20 expression and commonly express terminal B-cell differentiation markers such as MUM1/IRF4, but are often negative for CD79a, CD38, CD138 and EBER^9^. Molecular studies usually reveal a monoclonal IGH rearrangement and the absence of somatic mutations in the IGH variable regions^6^. The exact pathogenesis of HDN remains incompletely understood, but it is believed to involve a complex interplay between HHV8 infection, immune dysregulation, chronic inflammation, and genetic alterations^6, 13^. HHV8 is an oncogenic virus with the ability to manipulate various cellular pathways and evade immune responses through the expression of specific viral genes. Among these genes are viral interleukin-6 (vIL-6), latency-associated nuclear antigen (LANA), viral FLICE-inhibitory protein (vFLIP), and viral cyclin D, which collectively promote cell survival, proliferation, angiogenesis, cytokine production, and immune evasion^6, 14, 15^.

The diagnosis of HDN requires confirming HHV8 infection in the neoplastic cells, typically achieved through immunohistochemistry or molecular methods, as well as the exclusion of other HHV8-associated lymphoproliferative disorders^16^. Differential diagnosis involves distinguishing HDN from conditions such as primary effusion lymphoma (PEL), multicentric Castleman disease (MCD), germinotropic lymphoproliferative disorder (GLPD), plasmablastic lymphoma (PBL), and other types of diffuse large B-cell lymphoma (DLBCL) ^6, 9, 17, 18^. The prognosis of HDN is generally poor^19^. However, certain prognostic factors have been identified, including performance status, serum lactate dehydrogenase level, extranodal involvement, and HIV status^16^. Treatment of HDN presents challenges due to the absence of a standardized treatment regimen and poor response to conventional chemotherapy^19^. Treatment options often involve CODOX-M/IVAC or dose-adjusted EPOCH protocols^20^, whereas CHOP is generally considered inadequate due to relatively low complete response rates and short progression-free survival^8, 21^. Autologous or allogeneic stem cell transplantation may benefit certain patients, although the optimal timing and selection criteria remain unclear^22^. Ongoing research is exploring novel agents which target HHV8 or its associated pathways, including antiviral drugs, immunomodulatory drugs, monoclonal antibodies and small molecular inhibitors^23–25^. However, larger and randomized trials are needed to validate their efficacy and safety.

However, HDN remains poorly understood due to its rarity and diagnostic challenges. The aim of this study is to investigate the epidemiological characteristics of HDN and identify factors that influence the survival of patients with HDN based on a population-based study using the national Cancer Institute’s Surveillance, Epidemiology and End Results (SEER) database.

## Materials and Methods

### Data collection of HDN

Data for the current study were collected from the Surveillance, Epidemiology, and End Results (SEER) Program (https://seer.cancer.gov/), which is a comprehensive cancer database maintained by the esteemed National Cancer Institute (NCI) ^26^. The data retrieval process was facilitated using the SEER*Stat software version 8.4.0.6 (https://seer.cancer.gov/seerstat/, accessed on June 1, 2023) ^27^. Specifically, the "Incidence-SEER Research Plus Data, 17 Registries, Nov 2022 Sub (2000-2020)" was employed to identify patients diagnosed with HDN between 2011 and 2020 using the case listing session. The selection of cases was based on strict criteria, including the availability of known age (up to 89 years) and confirmation of malignant behavior. A flow chart (Figure 1) provided a visual representation of the inclusion and exclusion criteria applied. Only patients meeting the following inclusion criteria were considered: (1) histologic code according to the International Classification of Diseases for Oncology (ICD-O-3) (9738/3); (2) survival time was not 0 or unknown. Patients meeting any of the exclusion criteria were not included: (1) the type of reporting source was “laboratory only” or “other hospital outpatient unit or surgery center”; (2) the diagnosis confirmation was unknown or “clinical diagnosis only”. Ultimately, a final cohort of 67 patients with HDN was included in the analysis. Given that the SEER data used in this study are publicly available and the privacy of patients was duly protected through anonymization, ethical approval from an ethics committee was not deemed necessary.

**Figure 1.**
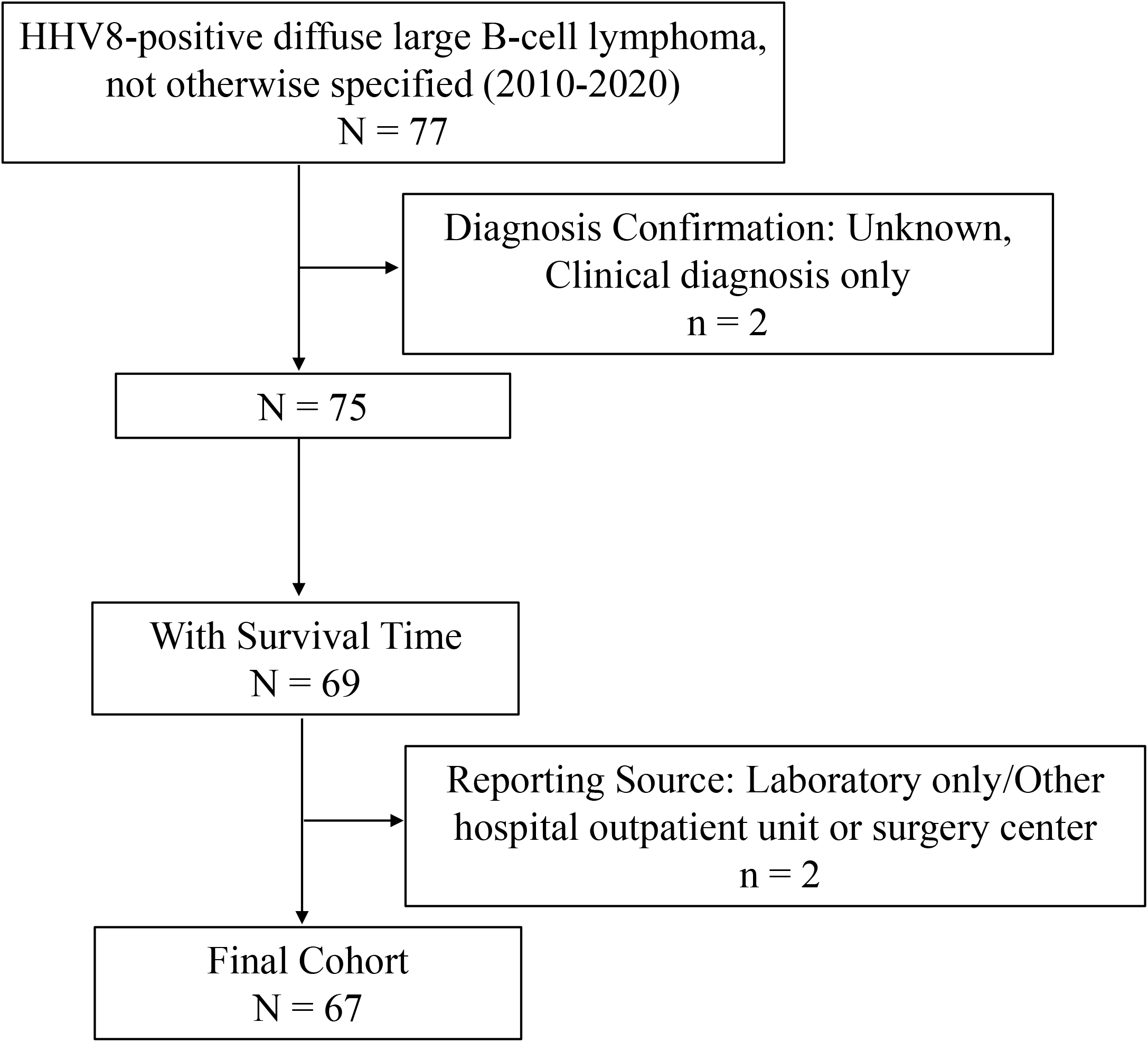
Flow chart of study cohort selection using the SEER database. A flow diagram of HDN patient selection in this study. HDN, HHV8-positive diffuse large B-cell lymphoma, not otherwise specified; SEER, Surveillance, Epidemiology, and End Results.

### Variable Definition of HDN

The variables collected for analysis included age, sex, race, marital status, year of diagnosis, primary site, vital status, survival months, COD to site recode, cause-specific death classification, cause of death to site, primary site, sequence number, first malignant primary indicator, total number of in situ/malignant tumors for patient, B symptoms, months from diagnosis to treatment, type of reporting source, diagnostic confirmation, chemotherapy recode, and radiation recode. Age was categorized into two groups: < 60 years old and 60 + years old, based on the age at diagnosis of HDN. Race were classified into White and Other (African American, Asian/Pacific Islander, American Indian/Alaska Native, and Unknown). Marital status was grouped into married, single, and other (including “Divorced”, “Separated”, “Widowed”, “Unmarried or domestic partner” and “Unknown”). Diagnosis year was grouped as 2011-2015 and 2016-2020. B symptoms referred to systemic symptoms of fever, night sweats, and weight loss that can occur with HDN. B symptoms were classified as “Yes” and “No/Unknown”. The variable “sequence number” was classified into two groups: primary HDN and secondary HDN (arises as a consequence of other primary malignancies). Treatment delay was derived from “months from diagnosis to treatment” and was grouped into 0, 1-7 and unknown (indicating “months from diagnosis to treatment” is unknown or > 24). Cause-of-death information was taken from the “COD to site recode” field. HDN-related death was defined in the SEER database as “dead (attributed to this cancer diagnosis)”. The definition of HDN-unrelated death was defined in the SEER database as death “dead (attributable to causes other than this cancer diagnosis)”. Overall survival time was calculated from the date of HDN diagnosis to death or last follow up.

### Statistical analysis

All statistical analyses in this study were conducted using the R programming language (version 4.2.1; R Foundation for Statistical Computing, Vienna, Austria). Student’s *t*-test and chi-square test were used to compare continuous and categorical data, respectively. Overall survival (OS) and disease-specific survival (DSS) were estimated using the Kaplan-Meier method with log-rank tests. Univariate and multivariate Cox proportional regression analyses were performed to determine the independent prognostic factors of HDN patients. Hazard ratios (HR) and 95% confidence intervals (95% CIs) were reported. Proportional hazards assumption tests and Schoenfeld residual tests were conducted to preliminarily evaluate the Cox proportional hazards regression model, and no violation of the proportion assumption was found for all variables. All *P*-values were two-sided, and a *P*-value < 0.05 was considered statistically significant.

## Results

### Baseline Characteristics of HDN Patients

A total of 67 patients were finally identified as HDN in the SEER 17 registry, Nov 2022 Sub (2000-2020) from January 2011 to December 2020, as depicted in Figure 1. Among these patients, lymph nodes were the most commonly affected sites (*n*=53, 79.1%), while extranodal sites (*n*=14, 20.9%) such as bone marrow, brain, cecum and small intestine were rarely affected. A detailed description of the primary site distribution of HDN was shown in Table 1. Within the study cohort, the majority of patients were male (79.1%), more than three folds that of female (*n*=14, 20.9%; Table 2). The average age at diagnosis was 51.8 ± 17.7 years, with 68.7% of patients being under 60 years old (< 60) and 31.3% being 60 years old or older (60+). The majority of HDN patients identified in this study were of White ethnicity (65.7%), while the remaining 34.3% comprised individuals from other ethnic backgrounds (including African American, Asian/Pacific Islander, American Indian/Alaska Native, and Unknown). Regarding marital status, 35.8% of patients were married, 32.8% had other marital statuses (such as divorced, widowed, separated, unmarried, or domestic partner), and 31.3% were single who had never been married. The incidence of HDN cases was 55.2% for the years 2011-2015 and 44.8% for 2016-2020. B symptoms (fever, night sweats, weight loss) were present in 44.8% of HDN patients. In terms of HDN classification, 76.1% of cases were categorized as primary HDN, while 23.9% were secondary HDN, arising as a result of other primary malignancies. Radiotherapy was received by a minimal number of patients (3.0%), whereas 55.2% underwent chemotherapy. Approximately 43.3% of patients received timely treatment, while 56.7% experienced delays of 1-7 months or the timing was unknown. At the time of the last follow-up, 41 (61.2%) patients were alive, 19 (28.4%) deaths were attributed to HDN, and an additional 7 (10.4%) patients died due to other causes, such as heart disease, kidney disease, accidents, and so on. The epidemiologic characteristics and survival comparison were summarized in Table 2.

**Table 1.** Distribution of the primary sites of HHV8-positive diffuse large B-cell lymphoma, not otherwise specified.

| Primary Sites | All Patients<br>(N=67) |
| --- | --- |
| **Extranodal (N=14, 20.9%)** |  |
| Bone marrow | 2 (3.0%) |
| Brain, NOS | 2 (3.0%) |
| Cardia, NOS | 1 (1.5%) |
| Cecum | 1 (1.5%) |
| Descended testis | 1 (1.5%) |
| Long bones of lower limb and associated joints | 1 (1.5%) |
| Mediastinum, NOS | 1 (1.5%) |
| Overlapping lesion of brain | 1 (1.5%) |
| Peritoneum, NOS | 1 (1.5%) |
| Pleura, NOS | 1 (1.5%) |
| Small intestine, NOS | 2 (3.0%) |
| **Nodal (N=53, 79.1%)** |  |
| abdominal lymph nodes | 1 (1.5%) |
| Intrathoracic lymph nodes | 1 (1.5%) |
| Lymph node, NOS | 12 (17.9%) |
| Lymph nodes of axilla or arm | 2 (3.0%) |
| Lymph nodes of head, face & neck | 4 (6.0%) |
| Lymph nodes of inguinal region or leg | 5 (7.5%) |
| Lymph nodes of multiple regions | 27 (40.3%) |
| Spleen | 1 (1.5%) |

Abbreviations: NOS, not otherwise specified.

**Table 2.** Baseline characteristics of patients with HDN.

| Characteristics | Total<br>(N=67) | Alive<br>(N=41) | Death |  |
| --- | --- | --- | --- | --- |
|  |  |  | HDN-related Death<sup>8</sup><br>(N=19) | Other cause<sup>9</sup><br>(N=7) |
| Sex, *n* (%) |  |  |  |  |
| Male | 53 (79.1%) | 30 (73.2%) | 17 (89.5%) | 6 (85.7%) |
| Female | 14 (20.9%) | 11 (26.8%) | 2 (10.5%) | 1 (14.3%) |
| Age, *n* (%) |  |  |  |  |
| <60 | 46 (68.7%) | 33 (80.5%) | 8 (42.1%) | 5 (71.4%) |
| 60+ | 21 (31.3%) | 8 (19.5%) | 11 (57.9%) | 2 (28.6%) |
| Race, *n* (%) |  |  |  |  |
| White | 44 (65.7%) | 28 (68.3%) | 13 (68.4%) | 3 (42.9%) |
| Other<sup>1</sup> | 23 (34.3%) | 13 (31.7%) | 6 (31.6%) | 4 (57.1%) |
| Marital Status, *n* (%) |  |  |  |  |
| Married | 24 (35.8%) | 18 (43.9%) | 5 (26.3%) | 1 (14.3%) |
| Single<sup>2</sup> | 21 (31.3%) | 11 (26.8%) | 6 (31.6%) | 4 (57.1%) |
| Other<sup>3</sup> | 22 (32.8%) | 12 (29.3%) | 8 (42.1%) | 2 (28.6%) |
| Diagnosis Year, *n* (%) |  |  |  |  |
| 2011-2015 | 37 (55.2%) | 25 (61.0%) | 8 (42.1%) | 4 (57.1%) |
| 2016-2020 | 30 (44.8%) | 16 (39.0%) | 11 (57.9%) | 3 (42.9%) |
| Site Classification, *n* (%) |  |  |  |  |
| Nodal | 53 (79.1%) | 34 (82.9%) | 12 (63.2%) | 7 (100%) |
| Extranodal | 14 (20.9%) | 7 (17.1%) | 7 (36.8%) | 0 (0%) |
| B Symptoms, *n* (%) |  |  |  |  |
| Yes | 30 (44.8%) | 19 (46.3%) | 7 (36.8%) | 4 (57.1%) |
| No/Unknown<sup>4</sup> | 37 (55.2%) | 22 (53.7%) | 12 (63.2%) | 3 (42.9%) |
| Sequence, *n* (%) |  |  |  |  |
| Primary HDN<sup>5</sup> | 51 (76.1%) | 31 (75.6%) | 13 (68.4%) | 7 (100%) |
| Secondary HDN<sup>6</sup> | 16 (23.9%) | 10 (24.4%) | 6 (31.6%) | 0 (0%) |
| RT, *n* (%) |  |  |  |  |
| Yes | 2 (3.0%) | 1 (2.4%) | 1 (5.3%) | 0 (0%) |
| No/Unknown | 65 (97.0%) | 40 (97.6%) | 18 (94.7%) | 7 (100%) |
|  |  |  | HDN-related Death<sup>8</sup><br>(N=19) | Other cause<sup>9</sup><br>(N=7) |
| Chemo, *n* (%) |  |  |  |  |
| Yes | 37 (55.2%) | 23 (56.1%) | 12 (63.2%) | 2 (28.6%) |
| No/Unknown | 30 (44.8%) | 18 (43.9%) | 7 (36.8%) | 5 (71.4%) |
| Treatment Delay, *n* (%) |  |  |  |  |
| 0 | 29 (43.3%) | 20 (48.8%) | 8 (42.1%) | 1 (14.3%) |
| 1-7/Unknown<sup>7</sup> | 38 (56.7%) | 21 (51.2%) | 11 (57.9%) | 6 (85.7%) |

<sup>1</sup> Races including American Indian, Asian/Pacific Islander, American Indian/Alaska Native and Unknown.
<sup>2</sup> Marital status at diagnosis was single (never married).
<sup>3</sup> Marital status of divorced, widowed, separated, unmarried or domestic partner and unknown at diagnosis.
<sup>4</sup> Indicated B symptoms not documented in medical record or not assessed or unknown if assessed.
<sup>5</sup> Sequence number of “primary only” and “1st of 2 or more primaries”, indicating HDN was the primary malignancy.
<sup>6</sup> Sequence number of “2nd of 2 or more primaries” and “3rd of 3 or more primaries”, indicating HDN was secondary to other primary malignancies.
<sup>7</sup> Unknown: months from diagnosis to treatment is unknown or > 24.
<sup>8</sup> Death attributable to HDN.
<sup>9</sup> Death of other cause such as diseases of heart, cerebrovascular diseases, septicemia, and so on.
Abbreviations: HDN, HHV8-positive diffuse large B-cell lymphoma, not otherwise specified; Chemo, chemotherapy; COD, cause of death; RT, radiation therapy.

### Survival Analysis of HDN Patients

During the follow-up period, there were 26 deaths, of which 19 were directly caused by the disease. As shown in Figure 2A and B, the 1-year, 3-year and 5-year OS was 67.4%, 65.6%, 58.4%, and 56.3%, respectively, and the corresponding DSS was 73.1%, 73.1%, and 67.8%, respectively. Notably, the group diagnosed between 2016-2020 exhibited significantly poorer overall survival compared to the group diagnosed between 2011-2015 (*P* = 0.040; Figure 2C). However, this significance did not persist for DSS (*P* = 0.074; Figure 2D). Moreover, the Kaplan-Meier analysis of overall survival revealed that only older age (*P* = 0.002, Figure 3A) was significantly associated with a lower overall survival rate. No significant differences were observed in overall survival based on sex, race, marital status, primary sites, sequence number, B symptoms, chemotherapy, or treatment delay categories (Figure 3B-I). Additionally, Figure 4 illustrates the Kaplan-Meier analysis of DSS, which showed similar results except for nodal sites, demonstrating significantly better survival compared to extranodal sites (*P* = 0.027, Figure 4E).

**Figure 2.**
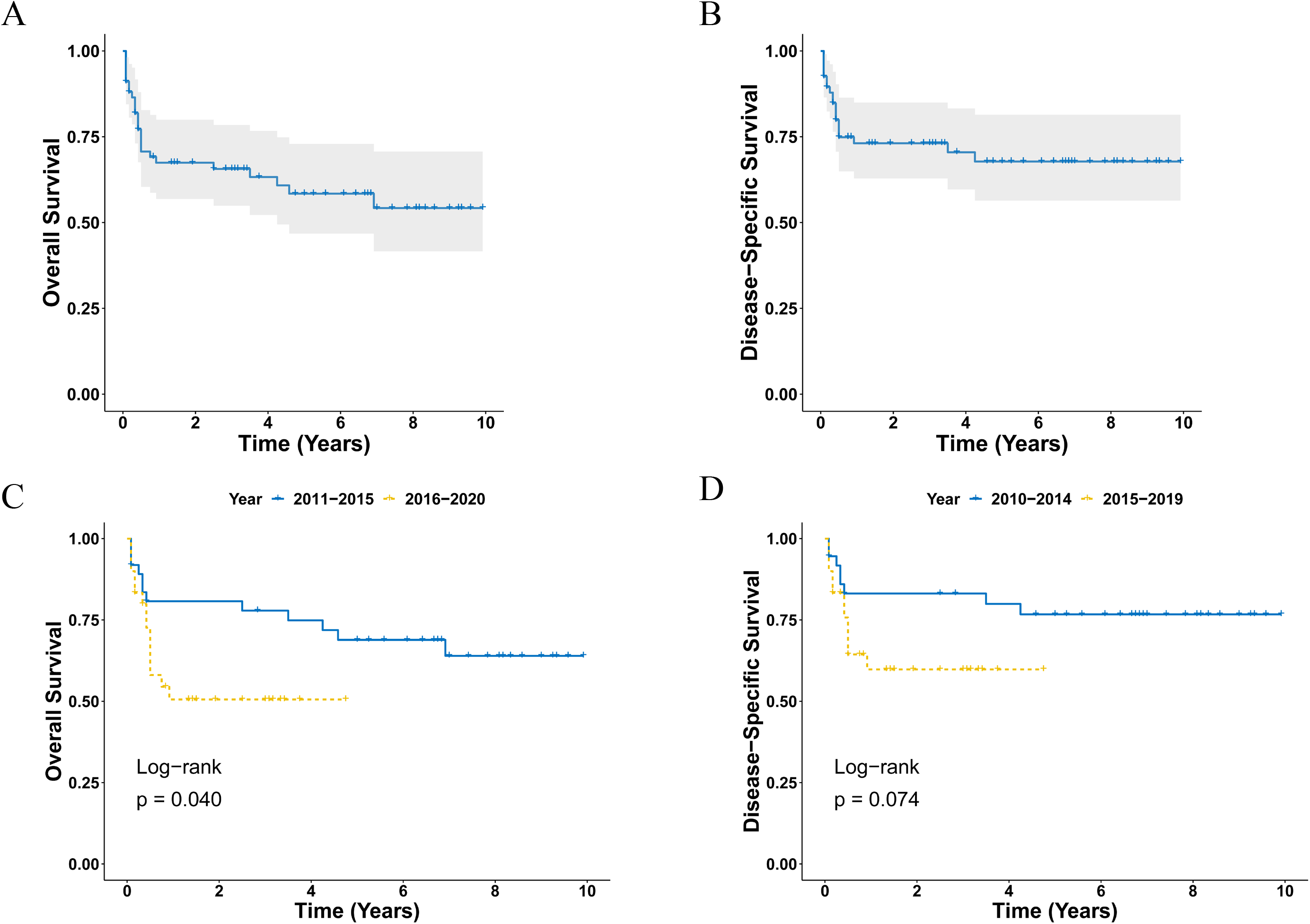
Survival analysis of primary HDN. **(A, B)** OS (A) and DSS (B) curves for all primary HDN patients. **(C, D)** Survival curves of OS (E) and DSS (F) according to the diagnosis years. HDN, HHV8-positive diffuse large B-cell lymphoma, not otherwise specified; OS, overall survival; DSS, disease-specific survival.

**Figure 3.**
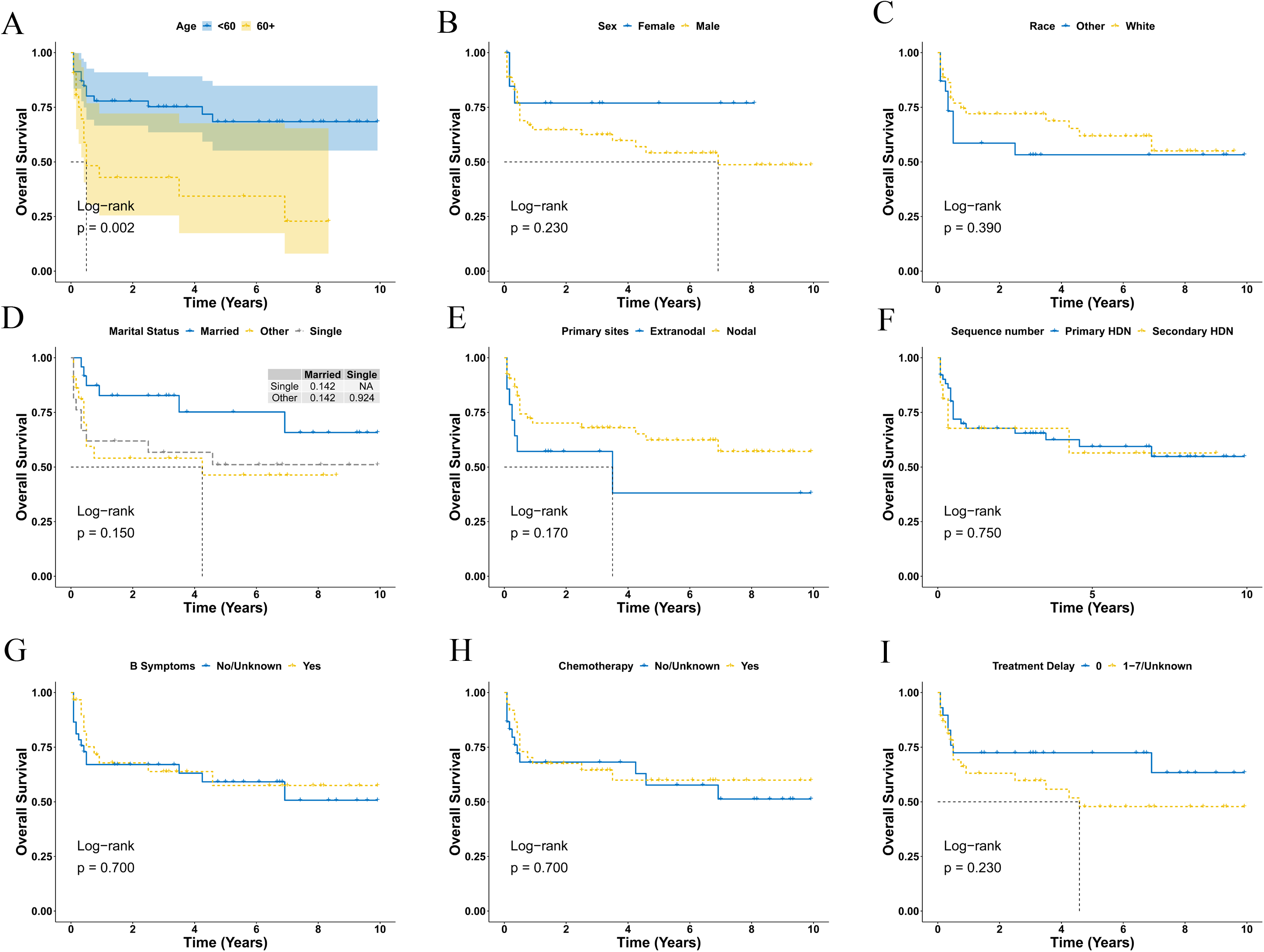
Overall survival analysis of primary HDN stratified by age (A), sex (B), race (C), marital status (D), primary sites (E), sequence number (F), B symptoms (G), chemotherapy (H) and treatment delay (I) using Kaplan-Meier method. HDN, HHV8-positive diffuse large B-cell lymphoma, not otherwise specified.

**Figure 4.**
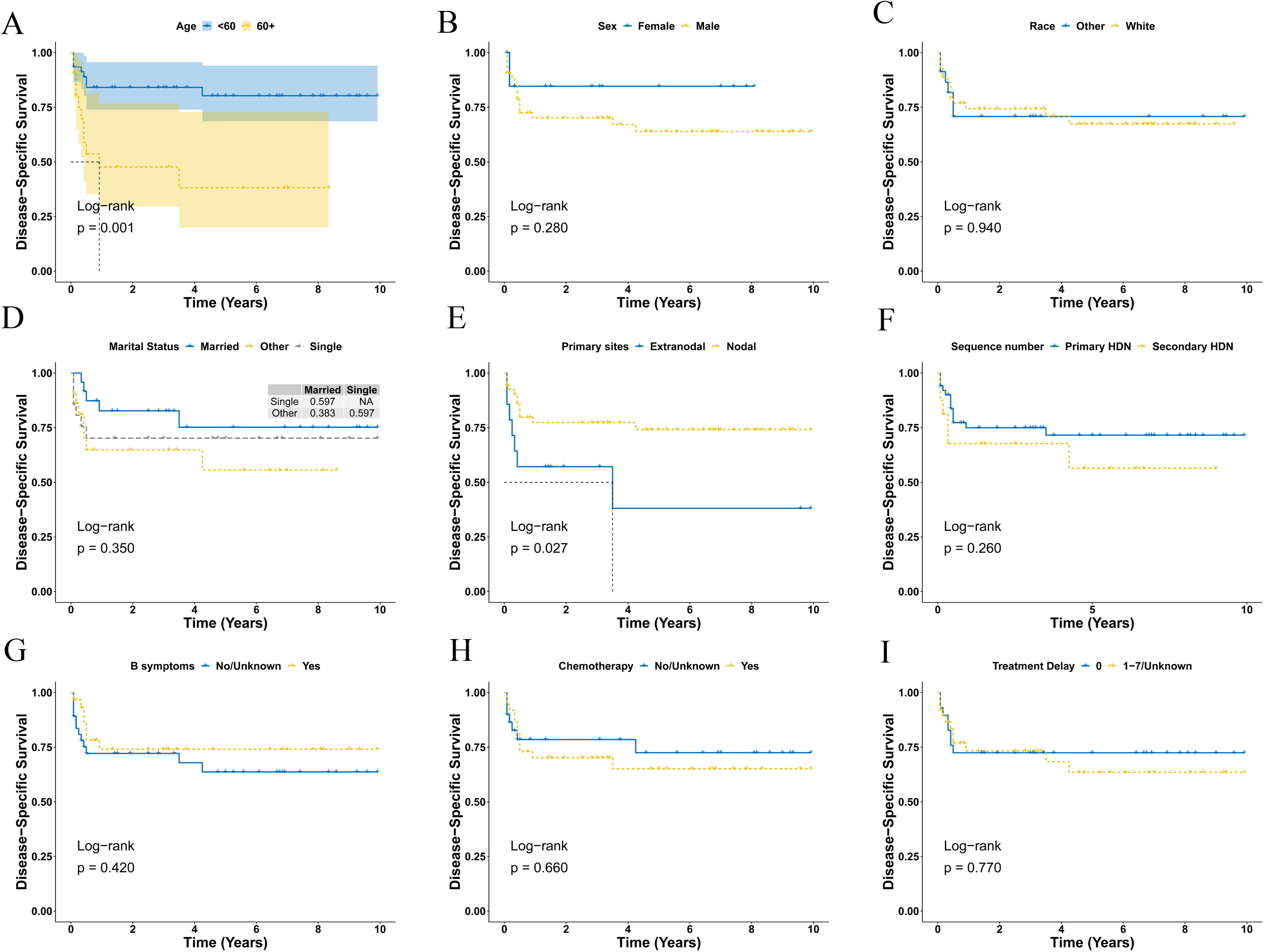
Disease-specific survival analysis of primary HDN stratified by age (A), sex (B), race (C), marital status (D), primary sites (E), sequence number (F), B symptoms (G), chemotherapy (H) and treatment delay (I) using Kaplan-Meier method. HDN, HHV8-positive diffuse large B-cell lymphoma, not otherwise specified.

### Univariate and Multivariable Cox Regression Analysis of HDN Patients

The univariate Cox regression analysis of OS showed that older age (HR: 3.12, CI= 1.44-678, *P* = 0.004) and being diagnosed between 2016-2020 (HR: 2.36, CI=1.02-5.49, *P* = 0.045) were significantly associated with poorer overall survival in patients with HDN (Table 3). However, variables such as sex, race, marital status, site classification, B symptoms, sequence, chemotherapy, and treatment delay did not demonstrate a significant impact on overall survival outcomes. When considering DSS, only age and site classification exhibited significant associations with worse survival (Table 3). Further analysis using multivariate Cox regression, as shown in Table 4, indicated that older age and being single as marital status were independent prognostic factors for OS. In the case of DSS, only age was identified as an independent prognostic factor (Table 4).

**Table 3.**
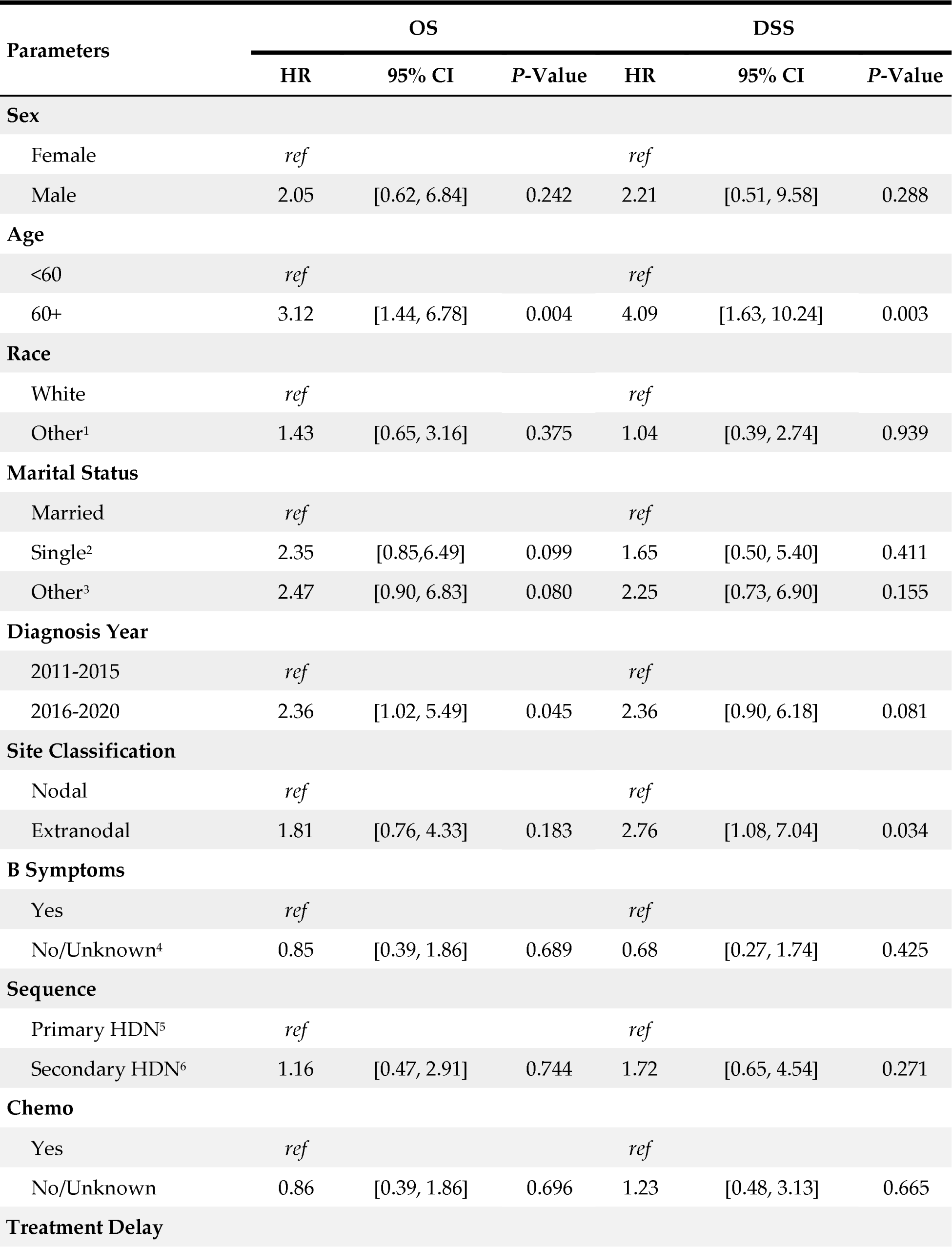
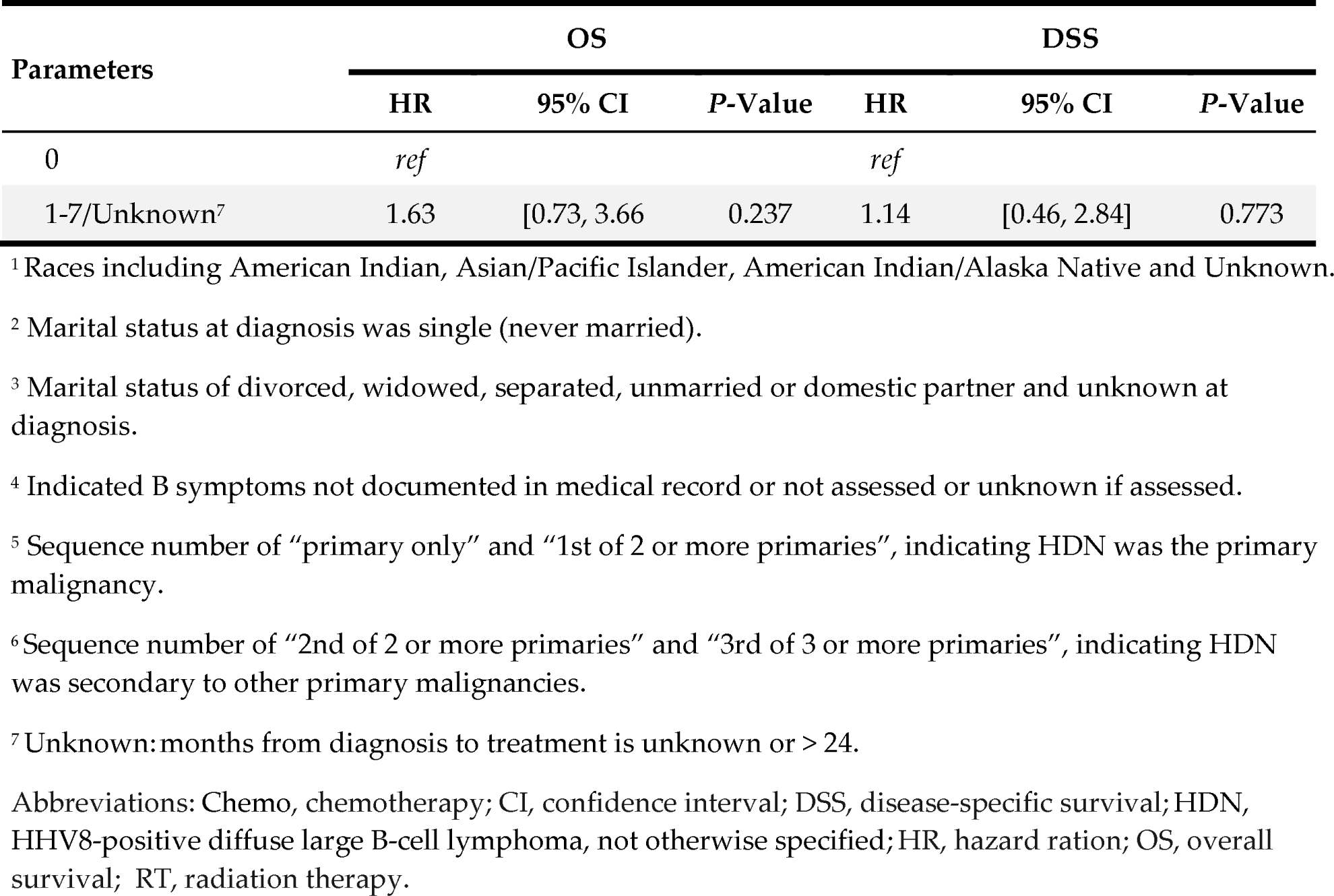
Univariable cox regression analysis for OS and DSS of HDN patients.

**Table 4.** Multivariable cox regression analysis for OS and DSS of HDN patients.

| Parameters | OS |  |  | DSS |  |  |
| --- | --- | --- | --- | --- | --- | --- |
|  | HR | 95% CI | P-Value | HR | 95% CI | P-Value |
| Sex |  |  |  |  |  |  |
| Female | ref |  |  | ref |  |  |
| Male | 3.36 | [0.89, 12.7] | 0.074 | 4.62 | [0.89, 24.10] | 0.069 |
| Age |  |  |  |  |  |  |
| <60 | ref |  |  | ref |  |  |
| 60+ | 6.54 | [2.06, 20.8] | 0.001 | 9.87 | [2.07, 47.20] | 0.004 |
| Race |  |  |  |  |  |  |
| White | ref |  |  | ref |  |  |
| Other<sup>1</sup> | 1.42 | [0.59, 3.40] | 0.429 | 1.00 | [0.35, 2.80] | 0.996 |
| Marital Status |  |  |  |  |  |  |
| Married | ref |  |  | ref |  |  |
| Single<sup>2</sup> | 5.93 | [1.64, 21.4] | 0.007 | 5.79 | [0.96, 34.90] | 0.055 |
| Other<sup>3</sup> | 2.25 | [0.64, 7.90] | 0.207 | 4.25 | [0.80, 22.60] | 0.089 |
| Diagnosis Year |  |  |  |  |  |  |
| 2011-2015 | ref |  |  | ref |  |  |
| 2016-2020 | 2.08 | [0.84, 5.20] | 0.113 | 1.64 | [0.59, 4.60] | 0.347 |
| Site Classification |  |  |  |  |  |  |
| Nodal | ref |  |  | ref |  |  |
| Extranodal | 1.79 | [0.61, 5.30] | 0.292 | 2.80 | [0.82, 9.60] | 0.102 |
| B Symptoms |  |  |  |  |  |  |
| Yes | ref |  |  | ref |  |  |
| No/Unknown<sup>4</sup> | 1.41 | [0.50, 4.00] | 0.510 | 1.69 | [0.47, 6.10] | 0.423 |
| Sequence |  |  |  |  |  |  |
| Primary HDN<sup>5</sup> | ref |  |  | ref |  |  |
| Secondary HDN<sup>6</sup> | 0.96 | [0.34, 2.70] | 0.945 | 1.57 | [0.54, 4.60] | 0.409 |
| Chemo |  |  |  |  |  |  |
| Yes | ref |  |  | ref |  |  |
| No/Unknown | 0.83 | [0.31, 2.20] | 0.707 | 2.07 | [0.48, 8.80] | 0.327 |
| Treatment Delay |  |  |  |  |  |  |
|  | HR | 95% CI | P-Value | HR | 95% CI | P-Value |
| 0 | *ref* |  |  | *ref* |  |  |
| 1-7/Unknown<sup>7</sup> | 1.34 | [0.54, 3.60] | 0.558 | 0.97 | [0.31, 3.10] | 0.965 |

<sup>1</sup> Races including American Indian, Asian/Pacific Islander, American Indian/Alaska Native and Unknown.
<sup>2</sup> Marital status at diagnosis was single (never married).
<sup>3</sup> Marital status of divorced, widowed, separated, unmarried or domestic partner and unknown at diagnosis.
<sup>4</sup> Indicated B symptoms not documented in medical record or not assessed or unknown if assessed.
<sup>5</sup> Sequence number of “primary only” and “1st of 2 or more primaries”, indicating HDN was the primary malignancy.
<sup>6</sup> Sequence number of “2nd of 2 or more primaries” and “3rd of 3 or more primaries”, indicating HDN was secondary to other primary malignancies.
<sup>7</sup> Unknown: months from diagnosis to treatment is unknown or > 24.
Abbreviations: Chemo, chemotherapy; CI, confidence interval; DSS, disease-specific survival; HDN, HHV8-positive diffuse large B-cell lymphoma, not otherwise specified; HR, hazard ratio; OS, overall survival; RT, radiation therapy.

## Discussion

In this study, we performed a population-based analysis of 67 patients with HDN, a rare and aggressive subtype of DLBCL that is associated with HHV8 infection. The results of this study provide a comprehensive overview of the epidemiology, treatment patterns and survival outcomes of HDN patients in the United States from 2011 to 2020. To our knowledge, this is the first population-based study to analyze the characteristics and prognosis of HDN patients using the SEER database. Our findings reveal some novel and important insights into this rare and aggressive lymphoma.

In this study, we found that HDN was predominantly a disease of males, with a male-to-female ratio of 3.8:1, which is consistent with previous reports^28^. The median age at diagnosis was 51.8 years, which is similar to that reported in other studies^8, 28^. We also observed that HDN had a higher incidence in whites than in other races, which is in agreement with previous findings^8^. However, the racial disparity in HDN incidence remains unclear and warrants further investigation. Moreover, we demonstrated that the primary site distribution of HDN was mainly nodal, accounting for 79.1% of all cases, while the extranodal sites such as bone marrow, brain, cecum, and small intestine were rarely involved. This is contrary to some previous studies that reported a higher frequency of extranodal involvement in HDN^28^. The reason for this discrepancy may be related to the differences in the inclusion criteria, sample size and geographic regions of the studies.

Moreover, we showed that the diagnosis year group of 2016-2020 had a significantly worse OS than the diagnosis year group of 2011-2015, but not for DSS. This finding suggests that there may be some changes in the natural history or treatment response of HDN over time. One possible explanation for this finding is that the newer cases may represent a more aggressive or resistant subtype of HDN that has emerged in recent years. Alternatively, the newer cases may reflect a different patient population that has more comorbidities or immunosuppression that affect their survival outcomes. Further studies with larger sample sizes and longer follow-up periods are needed to explore the reasons for this temporal difference in survival and to determine whether it is a real phenomenon or a statistical artifact. We also found that site classification was a significant prognostic factor for DSS in HDN patients, with nodal sites showing better survival than extranodal sites. This finding had not been reported by other researchers previously. The reason may be related to the different biological characteristics or therapeutic responses of nodal and extranodal tumors.

Previous studies demonstrated that age was the most important predictor of survival in DLBCL patients^2, 29^. In this study, we identified age as independent prognostic factors for OS and DSS. These findings indicate that age is a prognostic factor of survival in HDN patients, regardless of other clinical variables. The possible mechanisms underlying the association between age and survival may include the decreased immune function, increased comorbidities and reduced tolerance to therapy in older patients^30^. We also identified marital status of being single as independent prognostic factors for OS. The association between marital status and survival may reflect the social support and psychological well-being of patients with different marital statuses^31–33^. Single patients may have less social support and more psychological distress than married patients, which may adversely affect their adherence to treatment and quality of life^32^.

Up to now, the optimal chemotherapy regimen for HDN patients is not well established^19^. In the present study, we found that chemotherapy was not associated with improved survival outcomes in HDN patients. This is in contrast to some previous studies that reported a survival benefit from chemotherapy in HDN^8, 22, 25^. The reason for this discrepancy may be due to the heterogeneity of chemotherapy regimens among different studies. Therefore, more studies are needed to explore the optimal chemotherapy regimen for HDN patients.

However, our study has some limitations that should be acknowledged. First, due to the rarity of HDN, our sample size was relatively small, which may limit the statistical power and generalizability of our results. Second, SEER database lacks some important clinical information, such as HHV8 viral load, immunophenotype, molecular subtype, treatment regimen, and response to therapy. These factors may have an impact on the survival outcomes of HDN patients and should be considered in future studies. Third, our study design was retrospective and observational, which introduces the possibility of confounding variables and bias in the analysis and interpretation of the data. Fourth, due to the lack of standardized diagnostic criteria and reporting methods for HDN, there may be some misclassification or underreporting of cases in the SEER database, which may introduce bias or error into our analysis.

## Conclusions

In conclusion, HDN is a rare and aggressive subtype of DLBCL that mainly affects males with lymph node involvement. The prognosis for HDN remains discouraging, with age and marital status of being single as independent prognostic indicators. Notably, the administration of chemotherapy did not yield significant improvements in survival rates. Our study provides a comprehensive description and analysis of HDN patients from the SEER database, which is the largest cohort reported to date. Nevertheless, further prospective investigations involving larger sample sizes and extended follow-up periods are necessary to validate our findings and gain deeper insights into the pathogenesis, diagnosis, and treatment modalities of HDN.

## Data Availability Statement

The data analyzed in this study are from the SEER database (https://seer.cancer.gov/) that are available to the public.

## Acknowledgments

The interpretation of the data is the sole responsibility of the author(s). The author(s) acknowledge the efforts of the National Cancer Institute and the Surveillance, Epidemiology, and End Results (SEER) Program tumor registries in the creation of the SEER database.

## Abbreviations

Chemo: chemotherapy;
CI: confidence interval;
COD: cause of death;
DSS: disease-specific survival;
HDN: HHV8-positive diffuse large B-cell lymphoma, not otherwise specified;
HR: hazard ration;
OS: overall survival;
RT: radiation therapy;
SEER: Surveillance, Epidemiology, and End Results;
SPMs: second primary malignancies;
NOS: not otherwise specified.

## References

1. Chang Y, Cesarman E Fau - Pessin MS, Pessin Ms Fau - Lee F, et al. Identification of herpesvirus-like DNA sequences in AIDS-associated Kaposi’s sarcoma. (0036-8075 (Print))

2. Sehn LH, Salles G. Diffuse Large B-Cell Lymphoma. New England Journal of Medicine. 2021;384(9):842–858. doi:10.1056/NEJMra2027612

3. Dupin N, Diss TL, Kellam P, et al. HHV-8 is associated with a plasmablastic variant of Castleman disease that is linked to HHV-8–positive plasmablastic lymphoma. Blood. 2000;95(4):1406–1412. doi:10.1182/blood.V95.4.1406.004k26_1406_1412

4. Oksenhendler E, Boulanger E, Galicier L, et al. High incidence of Kaposi sarcoma–associated herpesvirus–related non-Hodgkin lymphoma in patients with HIV infection and multicentric Castleman disease. Blood. 2002;99(7):2331–2336. doi:10.1182/blood.V99.7.2331

5. Carbone A, Gloghini A. KSHV/HHV8-associated lymphomas. British Journal of Haematology. 2008;140(1):13–24. doi:10.1111/j.1365-2141.2007.06879.x

6. Vega F, Miranda RN, Medeiros LJ. KSHV/HHV8-positive large B-cell lymphomas and associated diseases: a heterogeneous group of lymphoproliferative processes with significant clinicopathological overlap. Modern Pathology. 2020/01/01/ 2020;33(1):18–28. doi:10.1038/s41379-019-0365-y

7. Li M-F, Hsiao C-H, Chen Y-L, et al. Human herpesvirus 8-associated lymphoma mimicking cutaneous anaplastic large T-cell lymphoma in a patient with human immunodeficiency virus infection. Journal of Cutaneous Pathology. 2012;39(2):274–278. doi:10.1111/j.1600-0560.2011.01814.x

8. Qunaj L, Castillo JJ, Olszewski AJ. Survival of patients with CD20-negative variants of large B-cell lymphoma: an analysis of the National Cancer Data Base. Leukemia & Lymphoma. 2018/06/03 2018;59(6):1375–1383. doi:10.1080/10428194.2017.1387912

9. Chadburn A, Said J, Gratzinger D, et al. HHV8/KSHV-Positive Lymphoproliferative Disorders and the Spectrum of Plasmablastic and Plasma Cell Neoplasms: 2015 SH/EAHP Workshop Report—Part 3. American Journal of Clinical Pathology. 2017;147(2):171–187. doi:10.1093/ajcp/aqw218

10. Gonzalez-Farre B, Martinez D, Lopez-Guerra M, et al. HHV8-related lymphoid proliferations: a broad spectrum of lesions from reactive lymphoid hyperplasia to overt lymphoma. Modern Pathology. 2017;30(5):745–760. doi:10.1038/modpathol.2016.233

11. Van J, Dave AA, Schwartz M, Mitchell J. S2443 A Case of HHV-8 Diffuse Large B-Cell Lymphoma - Not Otherwise Specified With Liver Infiltration. Official journal of the American College of Gastroenterology | ACG. 2020;115

12. Fenu EM, Beaty MW, O’Neill TE, O’Neill SS. Cardiac Involvement by Human Herpesvirus 8-Positive Diffuse Large B-Cell Lymphoma: An Unusual Presentation in a Patient with Human Immunodeficiency Virus. Case Reports in Pathology. 2022/01/17 2022;2022:1298121. doi:10.1155/2022/1298121

13. Song JY, Dirnhofer S, Piris MA, Quintanilla-Martínez L, Pileri S, Campo E. Diffuse large B-cell lymphomas, not otherwise specified, and emerging entities. Virchows Archiv. 2023/01/01 2023;482(1):179–192. doi:10.1007/s00428-022-03466-6

14. Calabrò ML, Sarid R. HUMAN HERPESVIRUS 8 AND LYMPHOPROLIFERATIVE DISEASES. Mediterranean Journal of Hematology and Infectious Diseases. 2018;10:e2018061. doi:10.4084/mjhid.2018.061

15. Järviluoma A, Koopal S, Räsänen S, Mäkelä TP, Ojala PiM. KSHV viral cyclin binds to p27KIP1 in primary effusion lymphomas. Blood. 2004;104(10):3349–3354. doi:10.1182/blood-2004-05-1798

16. Courville EL, Sohani AR, Hasserjian RP, Zukerberg LR, Harris NL, Ferry JA. Diverse Clinicopathologic Features in Human Herpesvirus 8–Associated Lymphomas Lead to Diagnostic Problems. American Journal of Clinical Pathology. 2014;142(6):816–829. doi:10.1309/ajcpuli3w6wuggpy

17. Ruiz-Cordero R, Lewis J, Blieden C, et al. Unusual immunophenotypic variant of large B-cell lymphoma associated with HHV-8 and EBV in an HIV positive patient. Human Pathology: Case Reports. 2015/06/01/ 2015;2(2):49–54. doi:10.1016/j.ehpc.2015.02.002

18. Pan Z-G, Zhang Q-Y, Lu Z-B, et al. Extracavitary KSHV-associated Large B-Cell Lymphoma: A Distinct Entity or a Subtype of Primary Effusion Lymphoma? Study of 9 Cases and Review of an Additional 43 Cases. The American Journal of Surgical Pathology. 2012;36(8):1129–1140. doi:10.1097/PAS.0b013e31825b38ec

19. Bower M, Carbone A. KSHV/HHV8-Associated Lymphoproliferative Disorders: Lessons Learnt from People Living with HIV. Hemato. 2021;2(4):703–712.

20. Foster WR, Bischin A, Dorer R, Aboulafia DM. Human Herpesvirus Type 8-associated Large B-cell Lymphoma: A Nonserous Extracavitary Variant of Primary Effusion Lymphoma in an HIV-infected Man: A Case Report and Review of the Literature. Clinical Lymphoma, Myeloma and Leukemia. 2016;16(6):311–321. doi:10.1016/j.clml.2016.03.013

21. Group W, Bower M, Palfreeman A, et al. British HIV Association guidelines for HIV-associated malignancies 2014. HIV Medicine. 2014;15(S2):1–92. doi:10.1111/hiv.12136

22. Noy A. Optimizing treatment of HIV-associated lymphoma. Blood. 2019;134(17):1385–1394. doi:10.1182/blood-2018-01-791400

23. Casper C, Krantz EM, Corey L, et al. Valganciclovir for Suppression of Human Herpesvirus–8 Replication: A Randomized, Double-Blind, Placebo-Controlled, Crossover Trial. The Journal of Infectious Diseases. 2008;198(1):23–30. doi:10.1086/588820

24. Monini P, Carlini F, Stürzl M, et al. Alpha Interferon Inhibits Human Herpesvirus 8 (HHV-8) Reactivation in Primary Effusion Lymphoma Cells and Reduces HHV-8 Load in Cultured Peripheral Blood Mononuclear Cells. Journal of Virology. 1999;73(5):4029–4041. doi:doi:10.1128/jvi.73.5.4029-4041.1999

25. Casper C. New approaches to the treatment of human herpesvirus 8-associated disease. Reviews in Medical Virology. 2008;18(5):321–329. doi:10.1002/rmv.583

26. Surveillance, Epidemiology, and End Results (SEER) Program (www.seer.cancer.gov) SEER*Stat Database: Incidence - SEER Research Plus Data, 17 Registries, Nov 2022 Sub (2000-2020) - Linked To County Attributes - Time Dependent (1990-2021) Income/Rurality, 1969-2021 Counties, National Cancer Institute, DCCPS, Surveillance Research Program, released April 2023, based on the November 2022 submission.

27. Surveillance Research Program, National Cancer Institute SEER*Stat software (seer.cancer.gov/seerstat) version 8.4.0.

28. Sukswai N, Lyapichev K, Khoury JD, Medeiros LJ. Diffuse large B-cell lymphoma variants: an update. Pathology. 2020;52(1):53–67. doi:10.1016/j.pathol.2019.08.013

29. Abu Sabaa A, Mörth C, Hasselblom S, et al. Age is the most important predictor of survival in diffuse large B-cell lymphoma patients achieving event-free survival at 24 months: a Swedish population-based study. British Journal of Haematology. 2021;193(5):906–914. doi:10.1111/bjh.17206

30. Mohile SG, Dale W, Somerfield MR, Hurria A. Practical Assessment and Management of Vulnerabilities in Older Patients Receiving Chemotherapy: ASCO Guideline for Geriatric Oncology Summary. Journal of Oncology Practice. 2018/07/01 2018;14(7):442–446. doi:10.1200/JOP.18.00180

31. Aizer AA, Chen M-H, McCarthy EP, et al. Marital Status and Survival in Patients With Cancer. Journal of Clinical Oncology. 2013;31(31):3869–3876. doi:10.1200/jco.2013.49.6489

32. Ding Z, Yu D, Li H, Ding Y. Effects of marital status on overall and cancer-specific survival in laryngeal cancer patients: a population-based study. Scientific Reports. 2021/01/12 2021;11(1):723. doi:10.1038/s41598-020-80698-z

33. Kravdal Ø. The impact of marital status on cancer survival. Social Science & Medicine. 2001/02/01/ 2001;52(3):357–368. doi:10.1016/S0277-9536(00)00139-8

